# Benchmarking observational analyses before using them to address questions trials cannot answer: an application to coronary thrombus aspiration

**DOI:** 10.1101/2021.09.06.21263156

**Authors:** Anthony A. Matthews, Issa J. Dahabreh, Ole Fröbert, Bertil Lindahl, Stefan James, Maria Feychting, Tomas Jernberg, Anita Berglund, Miguel A. Hernán

## Abstract

To increase confidence in the use of observational analyses when addressing effectiveness questions beyond those addressed by randomized trials, one can first benchmark the observational analyses against existing trial results. We use Swedish registry data to emulate a target trial similar to the TASTE randomized trial, which found no difference in the risk of death or myocardial infarction by 1 year with or without thrombus aspiration among individuals with ST-elevation myocardial infarction. We benchmark the emulation against the trial at 1 year, then extend the emulation’s follow up to 3 years and estimate effects in subpopulations underrepresented in the trial. As in the TASTE trial, the observational analysis found no differences in risk of outcomes by 1 year between groups (risk difference 0.7 (−0.7,2.0) and -0.2 (−1.3,1.0) for death and myocardial infarction respectively), so benchmarking was considered successful. We additionally show no difference in risk of death or myocardial infarction by 3 years, or within subpopulations by 1 year. Benchmarking against an index trial before using observational analyses to answer questions beyond those the trial could address allowed us to explore whether the observational data can be trusted to deliver valid estimates of treatment effects.

## Background

Randomized trials are the preferred approach to estimate causal effects of clinical interventions. Randomized trials, however, cannot answer all clinically relevant causal questions, including those about long-term treatment effects or effects in individuals that do not enroll in trials. Analyses of observational databases are often used to complement the estimates of randomized trials, but observational analysis estimates may be confounded because differences in risk between treatment groups may be explained by differences between the individuals in each group rather than by the effect of treatment [1,2]. Therefore, causal analyses of observational data adjust for known and measured confounders, though there is no guarantee that such adjustment suffices to eliminate confounding bias [3].

One possible approach to increase confidence in observational effect estimates is benchmarking, that is, to demonstrate the observational analysis is able to replicate an index trial’s findings (e.g., effect on death by 1 year) before using the observational data to estimate what the index trial could not estimate (e.g., effect on death by 3 years if the index trial had a follow-up of 1 year, or the effect within subpopulations that were not well represented in the index trial) [4]. As an example, consider the Thrombus Aspiration in ST-Elevation myocardial infarction in Scandinavia (TASTE) randomized trial as our index trial. TASTE found no difference in the risk of death or myocardial infarction by 30 days or 1 year when comparing percutaneous coronary intervention with and without thrombus aspiration among individuals with ST-elevation myocardial infarction (STEMI) in the Nordic countries [5,6]. TASTE was designed to study the effects of thrombus aspiration by 1 year after baseline; analyses of observational data may be able to complement these results and make further inferences beyond those made by the TASTE trial. Trust in such observational analyses designed to ask a similar question as TASTE would be increased if they agreed with the 1-year trial results.

Successful agreement, however, requires sufficient adjustment for confounding; and it is possible that the structure of confounding varies before and after the publication of the trial, especially if that trial contributed to changes in the reasons for receiving treatment. Before TASTE there was evidence that thrombus aspiration improved coronary artery flow after percutaneous coronary intervention, but it was unknown whether it improved clinical endpoints such as mortality [7–9]. In Sweden, before TASTE, this uncertainty resulted in some centers implementing the routine use of thrombus aspiration, while others left it to the discretion of the operator. After TASTE found no beneficial effect of thrombus aspiration, in Sweden, routine thrombus aspiration was largely reserved for patients with large thrombi in a coronary artery [10].

Here, we use observational data from the national Swedish Web-Based System for Enhancement and Development of Evidence-Based Care in Heart Disease Evaluated According to Recommended Therapies (SWEDEHEART) registry, which is the same registry in which TASTE was nested, to emulate a target trial similar to TASTE. By carrying out the observational analysis in the same registry as the trial, we ensure the causal question is asked in the same health care setting. After evaluating if observational data before and after TASTE are comparable, we informally benchmark the observational analysis results against the trial results at 1 year, then extend follow up to 3 years and explore effects in subpopulations by 1 year.

## The index randomized trial: TASTE

### Trial design and analysis

TASTE was a multicenter, prospective, randomized, controlled, open-label clinical trial carried out between June 2010 and March 2013 [5,6]. In total, 31 percutaneous intervention centers recruited participants; 29 in Sweden, 1 in Iceland, and 1 in Denmark. SWEDEHEART was used to collect information for Swedish participants. Individuals were eligible for TASTE if percutaneous coronary intervention was planned for the treatment of acute STEMI (see Table 1 for additional criteria). Individuals who accepted the invitation to participate were randomly assigned to receive either percutaneous coronary intervention with or without thrombus aspiration. The primary end point was death by any cause within 30 days of percutaneous coronary intervention, and additional analyses explored death by any cause, re-hospitalization for myocardial infarction, and stent thrombosis with 1 year of percutaneous coronary intervention. Data on clinical end points were obtained from the Cause of Death and SWEDEHEART registries. The intention-to-treat analyses compared 1-year risk curves from Kaplan-Meier analyses and estimated the corresponding average hazard ratios from Cox proportional-hazards models.

**Table 1:** Description of TASTE randomized trial, target trial, and target trial emulation using SWEDEHEART registry.

| Protocol component | TASTE (index randomized trial) | Target trial | Target trial emulation using SWEDEHEART |
| --- | --- | --- | --- |
| **Eligibility criteria** | - Age 18 years or older between June 15th 2010 and March 25th 2013<br>- Diagnosis of ST-elevated myocardial infarction as defined by chest pain suggestive for myocardial ischemia for at least 30 minutes before hospital admission, time from onset of symptoms of less than 24 hours, and an ECG with new ST-segment elevation in two or more contiguous leads of greater than or equal to 0.2 mV in leads V2-V3 and/or greater than or equal to 0.1 mV in other leads or a probable new-onset left bundle branch block.<br>- Planned percutaneous coronary intervention in one of the 29 Swedish, one Icelandic, or one Danish coronary intervention centers<br>- Minimum of 50% stenosis in culprit artery by visual estimate<br>- Correspondence between ECG findings and culprit artery pathoanatomy<br>- Possibility to perform thrombus aspiration<br>- No emergency coronary artery bypass grafting<br>- No previous randomization in the TASTE trial<br>- Provided informed consent | Same as TASTE apart from:<br>- Study period September 4th 2007 to January 4<sup>th</sup> 2016, excluding 15th June 2010 to 25th March 2013, which was the period of recruitment for TASTE<br>- Only in the Swedish coronary intervention centers<br>- Possibility to perform thrombus aspiration not assessed<br>- Correspondence between ECG findings and culprit artery pathoanatomy not assessed<br>- Individuals excluded if died on same day as percutaneous coronary intervention | Same as target trial apart from:<br>- No informed consent asked, so not able to exclude those who would have not been asked or who would have declined participation if asked |
| **Treatment strategies** | (1) No thrombus aspiration followed by percutaneous coronary intervention: balloon dilatation, balloon dilatation and stenting or direct stenting to achieve antegrade flow. Post-dilatation of stents is optional.<br>(2) Thrombus aspiration followed by percutaneous coronary intervention: thrombus aspiration with an Export aspiration catheter (Medtronic Inc., Santa Rosa, USA). Continuous manual suction is performed using a proximal-to-distal approach, which is defined as active aspiration during initial passage of the lesion. In lesions that cannot initially be passed with the thrombus aspiration catheter it is permitted to dilate the lesion with an angioplasty balloon up to a maximal nominal diameter size of 2.0 mm and attempt to advance the thrombus aspiration catheter for a second time. After thrombus aspiration, percutaneous coronary intervention is done as described above. | Same as TASTE | Same as target trial |
| **Treatment assignment** | Individuals randomized to a treatment strategy (by center) | Individuals would be randomized to a treatment strategy and were aware of the assigned strategy. | Individuals assigned to the strategy that their data were compatible with. Assignment was treated as if randomized within levels of the following baseline covariates: age, sex, hospital, diabetes, body mass index, smoking, hyperlipidemia, hypertension, previous infarction, previous percutaneous coronary intervention, previous coronary artery bypass graft, stenosis class, proportion stenosis in culprit artery, angiography finding, heart rate, systolic blood pressure, diastolic blood pressure, thrombolysis, warfarin, aspirin, clopidogrel, prasugrel, heparin, low molecular weight heparin, bivalirudin, and Gp2b3a inhibitors. |
| **Outcomes** | - Death from any cause<br>- Rehospitalization for myocardial infarction<br>- Stent thrombosis | Same as TASTE | Same as target trial apart from:<br>- No Stent thrombosis due to few events<br>Outcomes identified as following:<br>- Death from any cause from the Swedish Cause of Death register by 1 year<br>- Myocardial infarction from the SWEDEHEART register by 1 year |
| **Follow-up** | Starts at treatment assignment and ends at date of first outcome (separately for analysis of each outcome), migration, or 1 year | Same as TASTE apart from:<br>- Unable to identify migration date<br>- Started from day after percutaneous coronary intervention<br>- Follow up to 3 years | Same as target trial |
| **Causal contrasts** | Intention to treat effect, per-protocol effect | Intention to treat effect, per-protocol effect | Observational analogue of the per-protocol effect |
| Statistical analysis | - Kaplan Meier plots<br>- Treatment differences were assessed with the use of the log-rank test and Cox regression. | - For intention to treat analyses, survival curves estimated using a pooled logistic regression outcome model with an indicator for assigned treatment group, a flexible time-varying intercept, product terms between treatment group and time, and standardization of the period specific cumulative probabilities. Comparison of risks via differences and ratios then estimated.<br>- Per-protocol analyses use the same technique as above, but restricted to individuals who received their assigned treatment, and with the inclusion of baseline covariates in the outcome models.<br>- Non-parametric bootstrapping with 200 samples used to calculate 95% confidence intervals. | Same per protocol analysis as target trial with adjustment for baseline covariates |

**Table 2:** Baseline characteristics of eligible individuals from an observational emulation of a target trial of thrombus aspiration vs. no thrombus aspiration, SWEDEHEART registry, 2007-2016.

|  | Thrombus aspiration | No thrombus aspiration |
| --- | --- | --- |
| **n** | 3462 | 14760 |
| **Age (yrs) (median [IQR])** | 66.0 [57.0, 74.0] | 68.0 [60.0, 77.0] |
| **Female (%)** | 887 (25.6) | 4422 (30.0) |
| **Hospital (%)** |  |  |
| Borås | 15 (0.4) | 143 (1.0) |
| Danderyd | 134 (3.9) | 363 (2.5) |
| Eskilstuna | 67 (1.9) | 398 (2.7) |
| Falun | 211 (6.1) | 638 (4.3) |
| Gävle | 333 (9.6) | 506 (3.4) |
| Halmstad | 13 (0.4) | 267 (1.8) |
| Helsingborg | 22 (0.6) | 90 (0.6) |
| Huddinge | 28 (0.8) | 131 (0.9) |
| Jönköping | 49 (1.4) | 679 (4.6) |
| Kalmar | 84 (2.4) | 566 (3.8) |
| Karlskrona | 165 (4.8) | 619 (4.2) |
| Karlstad | 65 (1.9) | 778 (5.3) |
| Karolinska Solna | 255 (7.4) | 1099 (7.4) |
| Kristianstad | 4 (0.1) | 144 (1.0) |
| Linköping | 251 (7.3) | 713 (4.8) |
| Lund | 785 (22.7) | 1929 (13.1) |
| Malmö | 32 (0.9) | 199 (1.3) |
| Sahlgrenska | 164 (4.7) | 1387 (9.4) |
| Skövde | 50 (1.4) | 487 (3.3) |
| St Görans | 42 (1.2) | 62 (0.4) |
| Sunderbyn | 19 (0.5) | 281 (1.9) |
| Sundsvall | 26 (0.8) | 214 (1.4) |
| SÖS | 170 (4.9) | 260 (1.8) |
| Trollhättan | 65 (1.9) | 328 (2.2) |
| Umeå | 33 (1.0) | 327 (2.2) |
| Uppsala | 160 (4.6) | 810 (5.5) |
| Västerås | 85 (2.5) | 283 (1.9) |
| Örebro | 125 (3.6) | 998 (6.8) |
| Östersund | 6 (0.2) | 47 (0.3) |
| Östra sjukhuset | 4 (0.1) | 14 (0.1) |
| **Stenosis class (%)** |  |  |
| A | 168 (4.9) | 928 (6.3) |
| B1 | 882 (25.5) | 4550 (30.8) |
| B2 | 1346 (38.9) | 6035 (40.9) |
| C | 1060 (30.6) | 3214 (21.8) |
| Other | 6 (0.2) | 33 (0.2) |
| **Stenosis in culprit artery (%)** |  |  |
| 50-69% | 34 (1.0) | 198 (1.3) |
| 70-89% | 118 (3.4) | 1081 (7.3) |
| 90-99% | 510 (14.7) | 4406 (29.9) |
| 100% | 2800 (80.9) | 9075 (61.5) |
| **Angiography finding (%)** |  |  |
| Normal | 2 (0.1) | 16 (0.1) |
| 1 vessel | 1957 (56.5) | 7260 (49.2) |
| 2 vessels | 931 (26.9) | 4271 (28.9) |
| 3 vessels | 450 (13.0) | 2537 (17.2) |
| Left main | 117 (3.4) | 659 (4.5) |
| Missing | 5 (0.1) | 17 (0.1) |
| **BMI (kg/m<sup>2</sup>) (median [IQR])** | 26.0 [24.0, 29.0] | 26.0 [24.0, 29.0] |
| Missing (%) | 839 (24.2) | 3642 (24.7) |
| **Smoking status (%)** |  |  |
| Never | 1157 (33.4) | 5596 (37.9) |
| Ex smoker (> 1 month) | 957 (27.6) | 4045 (27.4) |
| Current smoker | 1038 (30.0) | 3999 (27.1) |
| Missing | 310 (9.0) | 1120 (7.6) |
| **Diabetes (%)** | 428 (12.4) | 2292 (15.5) |
| **Hyperlipidemia treatment (%)** | 724 (20.9) | 3327 (22.5) |
| **Hypertension treatment (%)** | 1340 (38.7) | 6628 (44.9) |
| **Previous myocardial infarction (%)** | 426 (12.3) | 1996 (13.5) |
| **Previous percutaneous coronary intervention (%)** | 368 (10.6) | 1563 (10.6) |
| **Previous coronary artery bypass grafting (%)** | 64 (1.8) | 337 (2.3) |
| **Thrombolysis (%)** | 16 (0.5) | 54 (0.4) |
| **Warfarin (%)** | 72 (2.1) | 303 (2.1) |
| **Aspirin (%)** | 3341 (96.5) | 14347 (97.2) |
| **Clopidogrel or ticlopidine (%)** | 2141 (61.8) | 6357 (43.1) |
| **Prasugrel (%)** | 118 (3.4) | 623 (4.2) |
| **Heparin (%)** | 2796 (80.8) | 12648 (85.7) |
| **Low-molecular weight heparin (%)** | 311 (9.0) | 883 (6.0) |
| **Bivalirudin (%)** | 1729 (49.9) | 7081 (48.0) |
| **Glycoprotein IIb/IIIa inhibitors (%)** | 1457 (42.1) | 3906 (26.5) |
| **Heart rate (median [IQR])** | 74.0 [61.0, 87.0] | 75.0 [63.0, 88.0] |
| Missing (%) | 245 (7.1) | 784 (5.3) |
| **Systolic blood pressure (median [IQR])** | 138.0 [120.0, 157.0] | 141.0 [125.0, 160.0] |
| Missing (%) | 257 (7.4) | 837 (5.7) |
| **Diastolic blood pressure (median [IQR])** | 80.0 [70.0, 95.0] | 84.0 [72.0, 96.0] |
| Missing (%) | 457 (13.2) | 1475 (10.0) |

### Trial results

As published in the original TASTE paper, during the enrollment period there were 11956 individuals with STEMI, approximately 9420 individuals potentially eligible for enrollment in Sweden and Iceland (eligible individuals unknown in Denmark), and 7244 individuals randomized to a treatment arm in Sweden, Iceland, and Denmark; 3621 were assigned to thrombus aspiration and 3623 to no thrombus aspiration [6]. Appendix 1 shows the baseline characteristics and Table 3 shows the 1-year risks and average hazard ratios. The risk of each individual outcome did not differ between the treatment groups. The 1-year risk of death was 5.3% in individuals in the thrombus aspiration group and 5.6% in the no thrombus aspiration group, with a hazard ratio of 0.94 (95% confidence interval, 0.78, 1.15). The 1-year risk of myocardial infarction was 2.7% in both groups, with a hazard ratio of 0.97 (0.73, 1.28). Stent thrombosis was rare; the 1-year risk was 0.7% in the thrombus aspiration group and 0.9% in the no thrombus aspiration group, with a hazard ratio of 0.84 (0.50,1.40).

**Table 3:**
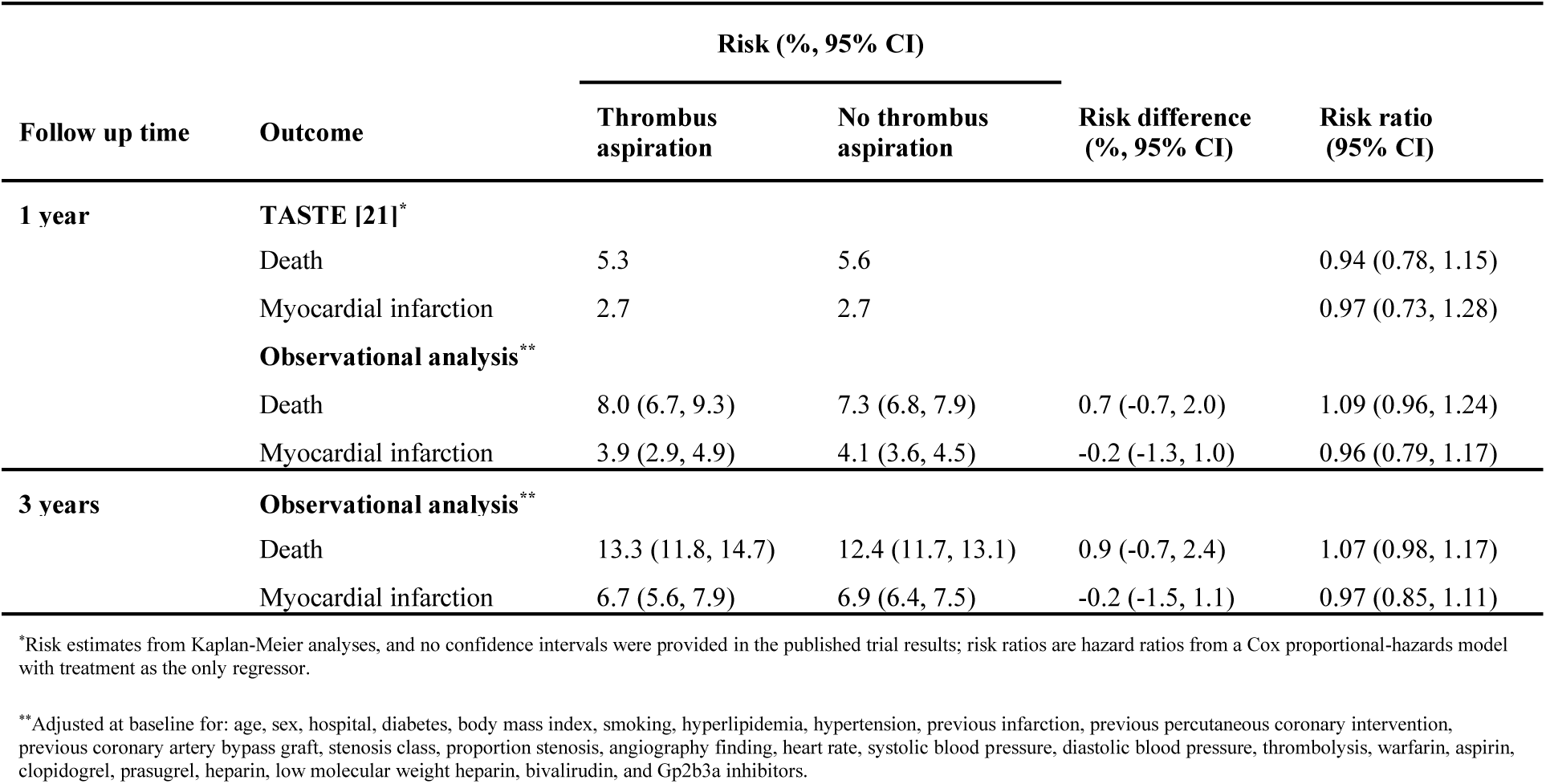
Estimated risks, risk differences, and risk ratios from the TASTE randomized trial and an observational emulation of a target trial of thrombus aspiration vs. no thrombus aspiration, SWEDEHEART registry, 2007-2016.

## The observational analysis

Causal inference from observational data can be seen as an attempt to emulate a pragmatic randomized trial—the target trial—that would answer the question of interest. The approach for emulating a target trial has two steps: 1) specify the protocol of the target trial and 2) emulate the target trial using the available observational data and appropriate methodology [11]. To compare TASTE to an observational analysis that aims to ask the same questions, using data from the SWEDEHEART registry, we first specified a protocol of a target trial similar to the protocol of TASTE, with deviations only when the observational data did not correspond to the information collected in the trial. We then emulated the target trial using the SWEDEHEART registry data. Table 1 summarizes all protocol elements from the target trial and its emulation, which we describe herein.

### The target trial protocol

#### Eligibility criteria

The eligibility criteria of the target trial would be the same as TASTE with five exceptions. First, the enrollment period would be September 2007 to January 2016, excluding June 2010 to March 2013, which is the period of participation in TASTE. Second, only Swedish coronary intervention centers would be included (no data are available from the Icelandic and Danish centers in the observational data). Third, possibility to perform thrombus aspiration would not be assessed. Fourth, correspondence between electrocardiogram findings and culprit artery pathoanatomy would not be assessed. Fifth, individuals that died on the day of percutaneous coronary intervention would be excluded and identification of outcomes would start from the day after percutaneous coronary intervention as it is not possible to distinguish if outcome events other than death, i.e., myocardial infarction, occurred before or after percutaneous coronary intervention when the events occurred on the same day as the procedure.

#### Treatment strategies

The treatment strategies in the target trial would be the same as those in TASTE: percutaneous coronary intervention (1) with thrombus aspiration, (2) without thrombus aspiration.

#### Treatment assignment

The target trial would randomly assign eligible individuals to one of the treatment strategies, and the physicians would be aware of the strategy to which the patient had been assigned.

#### Outcomes

The outcomes in the target trial would be death from any cause, myocardial infarction, or stent thrombosis.

#### Follow-up

The target trial would follow each individual from the day after treatment assignment until the outcome of interest (separate analysis for each outcome), or either 1 year for benchmarking or 3 years for analyses with extended follow up, whichever occurred first. It is not possible to identify migration date, so outcome data on those who migrated out of Sweden is unavailable. However, only about 0.5% of the Swedish population emigrates each year [12]. We expect this proportion to be even lower in individuals eligible for our study who recently had a myocardial infarction and are receiving regular healthcare.

#### Causal contrasts

The target trial would estimate the intention-to-treat effect, which is the effect of being assigned to thrombus aspiration or no thrombus aspiration, and the per-protocol effect, which is the effect of receiving the assigned thrombus aspiration or no thrombus aspiration.

#### Statistical analysis

For the intention-to-treat analysis, we estimate the survival curves in each group defined by assigned treatment strategy via a parametric pooled logistic model with an indicator for treatment group, a flexible time-varying intercept, and product terms between treatment group and time. We compare the estimated risks (one minus survival) via differences and ratios. To estimate the total effect on myocardial infarction, individuals who die are treated as not experiencing the outcome after death rather than as censored at death [13]. For the per-protocol analysis, we use the same technique as above, except that the analysis is restricted to individuals who received their assigned treatment, baseline covariates are included in the outcome models, and the estimated probabilities are standardized to the distribution of the baseline covariates [14]. Non-parametric bootstrapping with 200 samples is used to calculate 95% confidence intervals.

### Emulating the target trial in the SWEDEHEART registry

#### Data sources

SWEDEHEART collects data from all patients hospitalized for acute coronary syndrome or undergoing coronary or valvular intervention for any indication in all relevant hospitals across Sweden [15]. The registry was created by merging four existing cardiovascular healthcare quality registries in 2009: the Register of Information and Knowledge About Swedish Heart Intensive Care Admissions (RIKSHIA), the Swedish Coronary Angiography and Angioplasty Registry (SCAAR), the Swedish Heart Surgery Registry and the National Registry of Secondary Prevention (SEPHIA), and the Swedish Heart Surgery Registry. SWEDEHEART was used to collect information for patients when they were randomized in the TASTE trial, hence the data collection process was broadly similar between the two studies. SWEDEHEART is also linked to the Swedish National Patient Register, which records all primary and secondary diagnoses and procedures from inpatient hospitalizations and outpatient specialist care visits across Sweden; the Swedish Cause of Death register, which records all deaths and causes of death; and the Prescribed Drug register, which collects information on all dispensed medications [16–18].

#### Eligibility criteria

We identified individuals in the SWEDEHEART registry who met the eligibility criteria. As in all observational emulations, no informed consent was asked and hence we could not exclude individuals who would have not been asked or who would have declined participation if asked.

#### Treatment strategies and assignment

As treatment had already been given under routine clinical practice, we assigned eligible individuals in SWEDEHEART to the strategy their data were compatible with at baseline, and proceeded as if treatment was randomly assigned within levels of the following baseline covariates (full detail on covariates and their definitions in Appendix 2): age, sex, hospital, diabetes, body mass index, smoking, hyperlipidemia, hypertension, previous infarction, previous percutaneous coronary intervention, previous coronary artery bypass graft, stenosis class, proportion stenosis, angiography finding, heart rate, systolic blood pressure, diastolic blood pressure, thrombolysis, warfarin, aspirin, clopidogrel, prasugrel, heparin, low molecular weight heparin, bivalirudin, and glycoprotein 2b3a inhibitors.

#### Outcomes

We did not use stent thrombosis as an outcome because few events had been reported. We identified deaths from the Cause of Death register and myocardial infarctions from the SWEDEHEART registry. See Appendix 3 for further details on outcomes and their definitions.

#### Follow up

Follow up was the same as the target trial.

#### Causal contrasts

It was only possible to estimate the observational analog of the per-protocol effect as SWEDEHEART data collects information on the treatment an individual actually received, not what they were assigned.

#### Statistical analysis

The per-protocol analysis was the same as described above. Details of our modeling approach are presented in Appendix 4. We additionally stratified data by time period (before the TASTE trial began enrollment and after the TASTE trial completed enrollment), repeated the analyses, and compared estimates using data from each period to assess if results at 1 year of follow-up were comparable, regardless of changing reasons for receiving thrombus aspiration following publication of TASTE trial results.

We carried out nine sensitivity analyses: (1) we did not apply the eligibility criterion of 50% minimum stenosis (there was a high degree of missingness for the proportion stenosis variable used to identify this eligibility in the period before TASTE); (2) to understand the impact of measured covariates on effect estimates we conducted a separate analysis in which we adjusted for age and sex only, and we computed the difference between the fully-adjusted risk difference and the age and sex-adjusted risk difference; (3) we dropped all individuals with any missing data for baseline covariates (complete case analysis); (4) we censored individuals at death in the myocardial infarction analysis; (5) we defined myocardial infarction using a 2-day gap between discharge following the initial period in hospital and the new myocardial infarction event to account for individuals that were transferred between different hospitals without a new event; (6) we additionally included a Killip class variable in the models when data were stratified into time after TASTE (Killip class was collected from June 2009, so there was a high degree of missing data before TASTE); (7) we additionally included an indicator for time period (before or after the TASTE trial) in the models; (8) we estimated the standardized risk of each outcome separately in each treatment arm to allow for all possible interactions between treatment and covariates; and (9) we adjusted for baseline covariates using inverse probability weighting.

We informally benchmarked 1-year results from the emulation against the results of the TASTE trial. If risk contrasts when comparing those with and without thrombus aspiration were similar to those from TASTE, and the same clinical decision would be made regardless of the study used to inform the decision, benchmarking was deemed successful and analyses were replicated to estimate the 3-year risks, and data were stratified to estimate treatment effects by 1 year in subpopulations of individuals within stratums of sex (female/male), age (under 65/over 65), diabetes (no/yes), previous percutaneous coronary intervention (no/yes), and previous myocardial infarction (no/yes).

## Results

Figure 1 shows a flowchart of selection for the target trial emulation. There were 18222 eligible individuals, of whom 3462 were given thrombus aspiration and 14760 were not given thrombus aspiration. Table 2 shows the baseline characteristics of all eligible individuals. Before standardization, there were differences between groups for several variables including age, hospital, stenosis in culprit artery, and angiography finding (standardized mean differences in Appendix 5).

**Figure 1:**
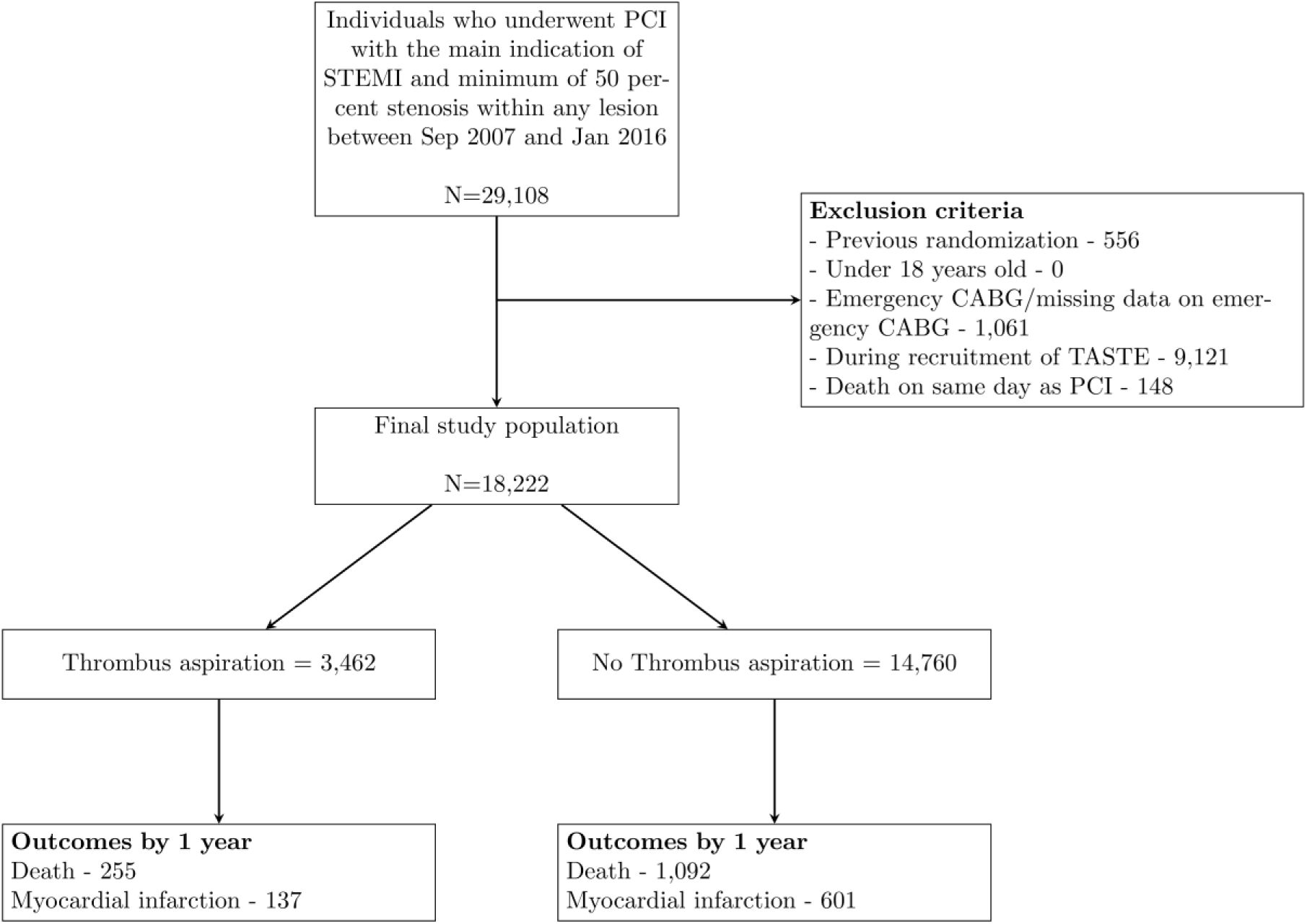
Flowchart of individuals eligible for an observational emulation of a target trial of thrombus aspiration vs. no thrombus aspiration, SWEDEHEART registry, 2007-2016.

Table 3 shows the estimated 1-year risks, risk differences, and risk ratios for death and myocardial infarction. The estimated risk (95% confidence interval) of death was 8.0% (6.7, 9.3) in the thrombus aspiration group and 7.3% (6.8, 7.9) in the group without thrombus aspiration; which results in a risk difference of 0.7% (−0.7, 2.0) and a risk ratio of 1.09 (0.96, 1.24). The estimated risk of myocardial infarction was 3.9% (2.9, 4.9) in the thrombus aspiration group and 4.1% (3.6, 4.5) in the group without thrombus aspiration; which results in a risk difference of -0.2% (−1.3, 1.0) and a risk ratio of 0.96 (0.79, 1.17).

Appendix 6 shows the baseline characteristics stratified by period (before and after TASTE enrollment), and Appendices 7 and 8 show results when using these stratified data for analysis. The 1-year risk of death and myocardial infarction did not differ between the treatment groups in both time periods, so use of data from both enrollment periods for benchmarking appears justified. Appendices 9-18 show results from sensitivity analyses; all results were broadly similar those from the primary analyses. Treatment groups were generally balanced in terms of the observed covariates after inverse probability weighting (Appendix 5).

### Benchmarking

Results of the target trial emulation at 1 year were informally benchmarked against results from the intention-to-treat analyses in TASTE (Table 3). Results appeared compatible within sampling variability: both the estimates from TASTE and the emulated target trial were very compatible with a similar range of hazard and risk ratio values for death (TASTE 95% CI: 0.78, 1.15; emulated target trial 95% CI: 0.96, 1.24) or myocardial infarction (TASTE 95%: 0.73, 1.28; emulated target trial 95% CI: 0.79, 1.17) by 1 year in the groups with or without thrombus aspiration.

### Extended follow up

Figure 2 shows the 3-year survival curves and Table 3 also shows the estimated 3-year risks, risk differences, and risk ratios. The estimated risk of death was 13.3% (11.8, 14.7) in the thrombus aspiration group and 12.4% (11.7, 13.1) in the group without thrombus aspiration, which results in a risk difference of 0.9% (−0.7, 2.4) and a risk ratio of 1.07 (0.98, 1.17). The estimated risk of myocardial infarction was 6.7% (5.6, 7.9) in the thrombus aspiration group and 6.9% (6.4, 7.5) in the group without thrombus aspiration; which results in a risk difference of -0.2% (−1.5, 1.1) and a risk ratio of 0.97 (0.85, 1.11).

**Figure 2:**
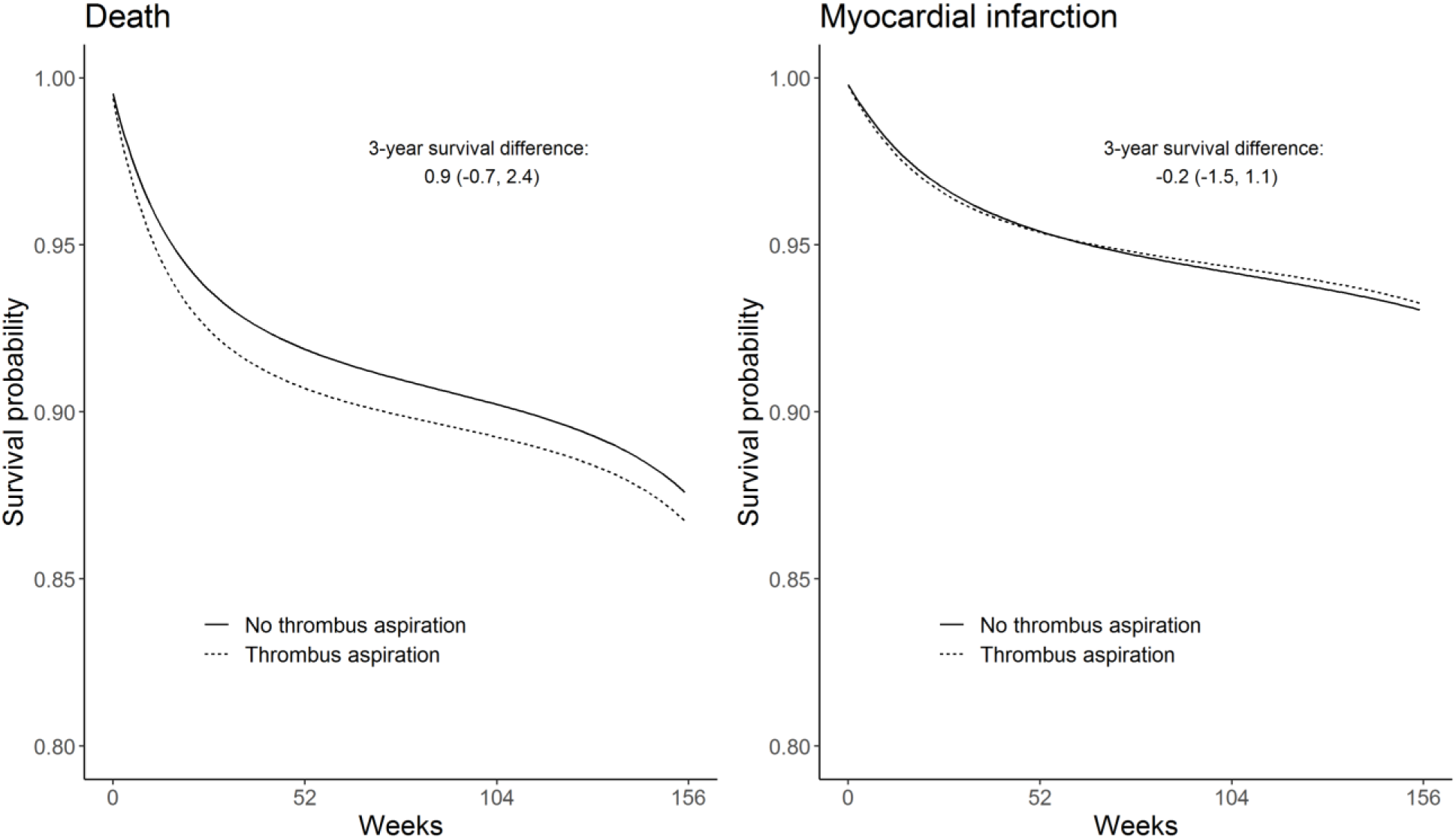
Standardized survival curves from an observational emulation of a target trial of thrombus aspiration vs. no thrombus aspiration, SWEDEHEART registry, 2007-2016.

### Subgroup effects

Table 4 shows the 1-year risks, risk differences, and risk ratios stratified by age, sex, diabetes, previous percutaneous coronary intervention, and previous myocardial infarction. Results were generally consistent with those from our main analyses.

**Table 4:** Estimated 1-year risks, risk differences, and risk ratios from an observational emulation of a target trial of thrombus aspiration vs. no thrombus aspiration, SWEDEHEART registry, 2007-2016, stratified by subpopulations.

|  | Risk (%; 95% CI) |  |  |  |
| --- | --- | --- | --- | --- |
| Outcome | Thrombus aspiration* | No thrombus aspiration* | Risk difference (%; 95% CI)* | Risk ratio (95% CI)* |
| SEX |  |  |  |  |
| Death |  |  |  |  |
| Female | 13.2 (10.8, 15.7) | 10.4 (9.3, 11.6) | 2.8 (-0.1, 5.6) | 1.27 (1.06, 1.51) |
| Male | 5.9 (4.6, 7.2) | 6.1 (5.5, 6.7) | -0.2 (-1.6, 1.2) | 0.96 (0.81, 1.15) |
| Myocardial infarction |  |  |  |  |
| Female | 4.4 (2.3, 6.5) | 4.1 (3.2, 4.9) | 0.3 (-2.0, 2.6) | 1.08 (0.75, 1.57) |
| Male | 3.7 (2.6, 4.8) | 4.1 (3.5, 4.7) | -0.4 (-1.7, 0.9) | 0.91 (0.72, 1.15) |
| AGE |  |  |  |  |
| Death |  |  |  |  |
| Over 65 | 12.0 (10.1, 13.9) | 10.9 (10.0, 11.8) | 1.1 (-0.9, 3.1) | 1.10 (0.97, 1.25) |
| Under 65 | 2.7 (1.4, 3.9) | 2.7 (2.2, 3.3) | -0.1 (-1.4, 1.3) | 0.98 (0.68, 1.41) |
| Myocardial infarction |  |  |  |  |
| Over 65 | 4.2 (2.8, 5.5) | 4.5 (3.8, 5.1) | -0.3 (-1.8, 1.2) | 0.93 (0.73, 1.19) |
| Under 65 | 3.5 (2.3, 4.8) | 3.5 (2.9, 4.2) | 0.0 (-1.4, 1.4) | 1.00 (0.76, 1.31) |
| DIABETES |  |  |  |  |
| Death |  |  |  |  |
| No | 7.5 (6.2, 8.8) | 6.4 (5.9, 7.0) | 1.0 (-0.4, 2.5) | 1.16 (1.01, 1.34) |
| Yes | 9.9 (6.2, 13.5) | 12.6 (10.9, 14.4) | -2.8 (-6.5, 1.0) | 0.78 (0.58, 1.05) |
| Myocardial infarction |  |  |  |  |
| No | 3.8 (2.8, 4.8) | 3.7 (3.2, 4.2) | 0.1 (-1.0, 1.3) | 1.03 (0.84, 1.27) |
| Yes | 3.9 (1.1, 6.7) | 6.1 (4.7, 7.4) | -2.2 (-5.3, 0.9) | 0.64 (0.36, 1.12) |
| PREVIOUS PERCUTANEOUS CORONARY INTERVENTION |  |  |  |  |
| Death |  |  |  |  |
| No | 8.0 (6.7, 9.3) | 7.4 (6.8, 8.0) | 0.6 (-0.8, 2.0) | 1.08 (0.95, 1.23) |
| Yes | 9.6 (5.6, 13.5) | 6.7 (5.0, 8.4) | 2.9 (-1.4, 7.2) | 1.43 (0.98, 2.07) |
| Myocardial infarction |  |  |  |  |
| No | 3.6 (2.5, 4.6) | 3.6 (3.2, 4.1) | -0.1 (-1.2, 1.1) | 0.98 (0.78, 1.22) |
| Yes | 6.8 (3.3, 10.3) | 7.7 (5.8, 9.6) | -0.9 (-4.9, 3.2) | 0.89 (0.58, 1.35) |
| PREVIOUS MYOCARDIAL INFARCTION |  |  |  |  |
| Death |  |  |  |  |
| No | 7.5 (6.3, 8.7) | 6.8 (6.2, 7.4) | 0.7 (-0.7, 2.1) | 1.10 (0.96, 1.27) |
| Yes | 12.1 (7.3, 16.8) | 11.0 (9.3, 12.6) | 1.1 (-3.9, 6.1) | 1.10 (0.80, 1.52) |
| Myocardial infarction |  |  |  |  |
| No | 3.8 (2.7, 4.9) | 3.5 (3.1, 4.0) | 0.3 (-0.9, 1.4) | 1.08 (0.87, 1.33) |
| Yes | 4.8 (1.9, 7.7) | 7.6 (6.0, 9.1) | -2.7 (-6.0, 0.6) | 0.64 (0.40, 1.02) |

\*Adjusted at baseline for: age, sex, hospital, diabetes, body mass index, smoking, hyperlipidemia, hypertension, previous infarction, previous percutaneous coronary intervention, previous coronary artery bypass graft, stenosis class, proportion stenosis, angiography finding, heart rate, systolic blood pressure, diastolic blood pressure, thrombolysis, warfarin, aspirin, clopidogrel, prasugrel, heparin, low molecular weight heparin, bivalirudin, and Gp2b3a inhibitors

## Discussion

We used observational data from the SWEDEHEART registry to address questions beyond those the TASTE trial could answer. The process had three steps. First, we used the observational data to emulate a target trial similar to TASTE, which estimated the effect of thrombus aspiration on risk of death and myocardial infarction by 1 year. Second, we informally benchmarked the observational analysis against TASTE by concluding that the same clinical decision would be made using either study because both studies estimated no benefit of thrombus aspiration. Third, in the observational analysis we extended follow up to 3 years to also estimate no benefit, and estimated no effects by 1 year in subpopulations defined by age, sex, diabetes, previous percutaneous coronary intervention, and previous myocardial infarction.

Unmeasured confounding is always a possibility in observational analyses. We were concerned that we could not adjust for thrombus burden, a predictor of myocardial infarction and death which affects the decision whether to administer thrombus aspiration, especially after TASTE when it was only used as a bail out for those with high thrombus burden in Sweden [10,19]. Lack of adjustment for this variable might explain the increased 1-year risk of death in the subpopulations of females and those without diabetes. However, a sensitivity analysis which additionally adjusted for Killip class, which is correlated with thrombus burden, did not considerably change the estimates (Appendix 15; analysis restricted to the time period after TASTE as Killip class was not available earlier) [20]. This implies either Killip class does not adequately capture thrombus burden or, more likely, there is little residual confounding due to thrombus burden.

Even in the absence of unmeasured confounding, there may be differences between the randomized trial and the observational analysis with respect to (1) study populations, (2) definition or measurement of interventions or outcomes, and (3) causal estimands. Because these differences may impact the estimates in different directions, it is logically possible that a partial cancelling out of these impacts leads to an erroneous conclusion that benchmarking was successful. We now discuss each of these differences and consider their impact on our results.

Between-study differences in effect estimates will occur if the treatment effect varies across groups that are unequally represented in each study. Eligible individuals who would not agree to enroll in a randomized trial is one such group, because those who do not enroll in trials (about 39% of individuals with STEMI in TASTE) are generally sicker with poorer prognosis. In TASTE, the 1-year risk of death was 5.3% in those who enrolled and were randomized to thrombus aspiration, and 16.4% in those who did not enroll and were given thrombus aspiration under routine practice. However, the inclusion of these individuals in the target trial emulation meant higher absolute risks than those who enrolled in TASTE, but this did not seem to affect the risk ratio estimates because among those not enrolled in TASTE the risks were similar when comparing groups with and without thrombus aspiration (16.4% and 15.7% for death, 3.8% and 3.7% for myocardial infarction) [6].

Another reason why study populations may differ is that observational data may not be detailed enough to match the eligibility criteria of the index trial. In our application, fewer individuals were eligible for the observational analysis in the period before TASTE compared with after TASTE, possibly because data on the proportion of stenosis in the culprit artery was less complete in SWEDEHEART in the earlier period (meaning the fewer people could be evaluated for the minimum of 50% stenosis criterion). However, in a sensitivity analysis which we did not use the minimum stenosis criterion to determine eligibility, effect estimates were broadly similar to the main results (Appendix 9).

Between-study differences in effect estimates will also occur if the measurement of interventions or outcomes varies between studies. However, this is unlikely to occur in our application because the randomized trial, TASTE, and the observational analysis were both embedded within the SWEDEHEART registry, and the definition and measurement of the intervention, thrombus aspiration, and the outcomes, death and myocardial infarction, were captured using the same mechanism. Use of the SWEDEHEART registry also means the health care system was the same in both studies.

Differences in causal estimands may also lead to between-study differences in effect estimates. In randomized trials, the main estimand is often the intention-to-treat effect, i.e., the effect of assignment to treatment. However, when using observational data, information may be only available on treatment an individual actually received, not what they were assigned or prescribed. Then, for point interventions like thrombus aspiration, the observational analysis can only estimate the per-protocol effect, i.e., the effect of receiving treatment [21]. Appropriate benchmarking then necessitates re-analyzing the randomized trial data to estimate the per-protocol effect, which requires adjustment for pre-randomization factors to account for confounding [22,23]. In our application, it is unlikely that differences in estimands affected the comparability of the estimates because adherence to the assigned treatment was very high (94%) in TASTE and, in fact, an unadjusted comparison restricted to the adherers resulted in a hazard ratio (0.95) very similar to that of the intention-to-treat analysis (0.94) [6].

Informal benchmarking at 1 year increases confidence in the reliability of observational inferences at 3 years and within sub-populations. Because increasing follow up increases the possibility of selection bias due to loss to follow-up, observational analyses generally require longitudinal data on joint predictors of loss to follow-up and the outcome interest. In our study, however, loss to follow-up is a minor concern because <0.5% of individuals emigrate each year [12]. We can also not think of baseline confounders that introduce bias only after 1 year. Additionally, our main analysis relies on the assumption that the measured covariates are approximately sufficient to adjust for confounding when estimating the effect in the entire study population. That is, we assume the magnitude of unmeasured confounding is, on average, small across all subgroups defined by the measured covariates. However, the magnitude of unmeasured confounding might be greater (or smaller) in certain subgroups and thus some subgroup-specific effect estimates may be more (or less) biased than the effect estimates in the entire study population.

We carried out an observational analysis that emulates a target trial, informally benchmarked its results with those from an index randomized trial, and used the observational analyses to draw causal inferences over a longer follow up duration and within sub-populations. The agreement between estimates from TASTE and our emulated target trial suggests that the observational data can deliver approximately valid estimates of treatment effects. This example shows how high-quality observational data can complement results from randomized trials and provide additional evidence to support clinical decision making.

## Data Availability

Pseudonymized personal data were obtained from national Swedish Registry holders after ethical approval and secrecy assessment. According to Swedish laws and regulations, personal sensitive data can only be made available for researchers who fulfill legal requirements for access to personal sensitive data.

## Declarations

### Funding

This work was supported by a grant from the Swedish Research Council (2018-03028). AM received funds from Strategic Research Program in Epidemiology at Karolinska Institutet and FORTE (2020-00029) during the conduct of the study. ID received funds from PCORI (ME-1502-27794) during the conduct of the study.

### Competing interests

AM, ID, BL, SJ, MF, TJ, and AB have nothing to disclose; OF reports grants from Sanofi Pasteur, during the conduct of the study; MH reports personal fees from Cytel and ProPublica, during the conduct of the study.

### Code availability

All analysis code is available at: https://github.com/tonymatthews/taste.

### Authors’ contributions

AM and MH designed the study, with assistance from ID and AB. AM carried out all analyses. AM drafted the manuscript. All authors contributed to further drafts and approved the final manuscript.

### Ethics approval

This study was approved by the Regional Ethical Review Board in Stockholm (2012/60-31/2)

## Notes

### Author Declarations

This study was approved by the Regional Ethical Review Board in Stockholm (2012/60-31/2)

### Summary of Updates

This is the latest update of the manuscript

