## appendix for "Benchmarking observational analyses before using them to address questions trials cannot answer: an application to coronary thrombus aspiration"

### APPENDICES

#### Appendix 1 – Baseline characteristics of the randomized patients enrolled in TASTE, according to treatment group

| Characteristic | No thrombus aspiration (N=3623) | Thrombus aspiration (N=3621) |
|---|---|---|
| Mean age (yr) $\pm$ SD | 65.9 $\pm$ 11.7 | 65.5 $\pm$ 11.5 |
| Male (%) | 2703 (74.6) | 2721 (75.1) |
| Diabetes (%) | 453 (12.5) | 448 (12.4) |
| Current smoking (%) | 1173 (32.4) | 1083 (29.9) |
| Previous myocardial infarction (%) | 440 (12.1) | 402 (11.1) |
| Previous PCI (%) | 362 (10.0) | 337 (9.3) |
| Previous CABG (%) | 74 (2.0) | 70 (1.9) |
| Fibrinolysis before PCI (%) | 69 (1.9) | 69 (1.9) |
| Procedure related medication (%) | | |
| Acetylsalicylic acid | 3542 (97.8) | 3546 (97.9) |
| Clopidogrel or ticlopidine | 2395 (66.1) | 2384 (65.8) |
| Ticagrelor | 1015 (28.0) | 1050 (29.0) |
| Prasugrel | 538 (14.8) | 562 (15.5) |
| Heparin | 3074 (84.8) | 3063 (84.6) |
| Low molecular-weight heparin | 142 (3.9) | 147 (4.1) |
| Bivalirudin | 2835 (78.3) | 2874 (79.4) |
| Glycoprotein IIb/IIIa inhibitor | 630 (17.4) | 558 (15.4) |
| Time from diagnostic ECG to PCI (min) | | |
| Median | 66 | 67 |
| $25^{th}$-$75^{th}$ percentile | 47-93 | 48-95 |
| Killip class $\geq$ 2 (%) | 183 (5.1) | 198 (5.5) |
| Radial-artery approach (%) | 2415 (66.7) | 2394 (66.1) |
| Type of disease (%) | | |
| One-vessel | 1940 (53.5) | 1946 (53.7) |
| Two-vessel | 1072 (29.6) | 1010 (27.9) |
| Three-vessel | 498 (13.7) | 558 (15.4) |
| Left main coronary artery disease | 105 (2.9) | 98 (2.7) |
| Data not available | 8 (0.2) | 9 (0.2) |
| TIMI flow grade 0 or 1 (%) | 2811 (77.6) | 2821 (77.9) |
| Thrombus grade (%) | | |
| G0 | 543 (15.0) | 490 (13.5) |
| G1 | 809 (22.3) | 733 (20.2) |
| G2 | 329 (9.1) | 341 (9.4) |
| G3 | 818 (22.6) | 887 (24.5) |
| G4 | 863 (23.8) | 903 (24.9) |
| G5 | 215 (5.9) | 235 (6.5) |
| Unknown | 46 (1.2) | 32 (0.9) |

SD = standard deviation; PCI = percutaneous coronary intervention; CABG = coronary artery bypass graft; ECG = electrocardiogram; min = minutes; TIMI = thrombolysis in myocardial infarction

### Appendix 2 - Covariate definitions and model specification in the target trial

| Covariate | Register used | Definition | Form | Categories |
|---|---|---|---|---|
| **Age** | SCAAR | | Linear, quadratic | N/A |
| **Gender** | SCAAR | | Indicator | Male/female |
| **PCI Centre** | SCAAR | Borås, Danderyd, Eskilstuna, Falun, Gävle, Halmstad, Helsingborg, Huddinge, Jönköping, Kalmar, Karlskrona, Karlstad, Karolinska Solna, Kristianstad, Linköping, Lund, Malmö, Sahlgrenska, Skövde, St Görans, Sunderbyn, Sundsvall, SÖS, Trollhättan, Umeå, Uppsala, Västerås, Örebro, Östersund, Östra sjukhuset | 30 categories | As in definition |
| **Stenosis class** | SCAAR | 1 - A<br>2 - B1<br>3 - B2<br>4 - C<br>5 - B1 Bifurcation<br>6 - B2 Bifurcation<br>7 - C Bifurcation<br>8 - Other<br>9 - Missing | 9 categories | 1, 2, 3, 4, 5, 6, 7, 8, missing |
| **Proportion stenosis** | SCAAR | Proportion stenosis in lesion with maximum stenosis filling<br>1 - 50-69%<br>2- 70-89%<br>3 - 90-99%<br>4 - 100% | 4 categories | 1, 2, 3, 4 |
| **Angiography finding** | SCAAR | 0 - normal<br>1 - 1 vessel + no left main<br>2 - 2 vessels + no left main<br>3 - 3 vessels + no left main<br>4 - left main | 5 categories | 0, 1, 2, 3, 4 |
| **Body mass index** | SCAAR | Weight/Height^2<br><br>In main analysis missing imputed with median. Any individuals with BMI above 50 set to 50, and individuals with BMI of below 10 set to 10 | Linear, quadratic | N/A |
| **Smoking** | SCAAR | 0 - never<br>1 - ex-smoker (> 1month)<br>2 - current<br>9 - missing | 4 categories | 0, 1, 2, missing |
| **Diabetes** | SCAAR | If missing assumed no | Indicator | Yes/No |
| **Prior myocardial infarction** | SCAAR | If missing assumed no | Indicator | Yes/No |
| **Prior percutaneous coronary intervention** | SCAAR | If missing assumed no | Indicator | Yes/No |
| **Prior coronary artery bypass grafting** | SCAAR | If missing assumed no | Indicator | Yes/No |
| **Prior treatment for hypertension** | SCAAR | If missing assumed no | Indicator | Yes/No |

| Covariate | Register used | Definition | Form | Categories |
|---|---|---|---|---|
| **Prior lipid lowering treatment** | SCAAR | If missing assumed no | Indicator | Yes/No |
| **Thrombolysis** | SCAAR | Before or under PCI, if missing assumed no | Indicator | Yes/No |
| **Warfarin** | SCAAR | Before or under PCI, if missing assumed no | Indicator | Yes/No |
| **Aspirin** | SCAAR | Before or under PCI, if missing assumed no | Indicator | Yes/No |
| **Clopidogrel/ticlopidine** | SCAAR | Before or under PCI, if missing assumed no | Indicator | Yes/No |
| **Prasugrel** | SCAAR | Before or under PCI, if missing assumed no | Indicator | Yes/No |
| **Heparin** | SCAAR | Before or under PCI, if missing assumed no | Indicator | Yes/No |
| **Low molecular weight heparin** | SCAAR | Before or under PCI, if missing assumed no | Indicator | Yes/No |
| **Bivalirudin** | SCAAR | Before or under PCI, if missing assumed no | Indicator | Yes/No |
| **GpIIb inhibitors** | SCAAR | Before or under PCI, if missing assumed no | Indicator | Yes/No |
| **Heart rate** | RIKSHIA | Some missing as captured from RIKSHIA and not all individuals have related RIKSHIA record.<br><br>In main analysis missing imputed with median | Linear, quadratic | N/A |
| **Systolic blood pressure** | RIKSHIA | Some missing as captured from RIKSHIA and not all individuals have related RIKSHIA record.<br><br>In main analysis missing imputed with median | Linear, quadratic | N/A |
| **Diastolic blood pressure** | RIKSHIA | Some missing as captured from RIKSHIA and not all individuals have related RIKSHIA record.<br><br>In main analysis missing imputed with median | Linear, quadratic | N/A |

SCAAR - Swedish Coronary Angiography and Angioplasty Registry; RIKSHIA - Registry of Information and Knowledge about Swedish Heart Intensive care Admissions; BMI = body mass index; PCI = percutaneous coronary intervention; GpIIb inhibitors = glycoprotein IIb/IIIa inhibitor

### Appendix 3 - Outcome definitions in the target trial

| TASTE trial | Target trial definition |
|---|---|
| **Death from any cause** | 1. SCAAR<br>The SCAAR register was used to identify date of death.<br><br>2. Cause of death register<br>If there was no death record in the SCAAR register, but a record of death in the cause of death registry, then date of death was taken from cause of death register. |
| **Myocardial infarction** | RIKSHIA<br>a. Between PCI and discharge date<br>If there was a record of reinfacrction related to the individuals record for intial myocardial infarction nd resuts percutaneous coronary intervention, then the date of new myocardial infarction was considered as the date of discharge. The actual date of reinfarction is not rcorded in SWEDEHEART, but the median time between original admission and discharge is 3 days.<br><br>b. After inital discharge date<br>After the individual for the intial period of care following percutaneous coronary intervetion through until the end of follow up, all myocardial infarction records in the RIKSHIA register were used (ICD 10 codes I21/I22). |

* SCARR - Swedish Coronary Angiography and Angioplasty Registry
** RIKSHIA - Registry of Information and Knowledge about Swedish Heart Intensive care Admissions

#### Appendix 4 - Standardization model specification

In the target trial the intention to treat effect is the effect of being assigned to thrombus aspiration followed by percutaneous coronary intervention versus percutaneous coronary intervention alone on the risk of death, myocardial infarction, or stent thrombosis. Estimating its observational analog requires adjustment for baseline confounders, which we achieved through standardization. First, we fit the pooled logistic regression model:

$$logit\left( Pr\left[ Y_{m+1} = 0 \mid A, L, Y_{m} = 0 \right] \right) = \beta_0 + \beta_1 m + \beta_2 m^2 + \beta_3 A + \beta_4 Am + \beta_5 Am^2 + \beta_6^T L$$

where $Y_{m+1}$ is the indicator for the outcome of interest at time $m + 1$ (observed among individuals who did not experience the outcome in the previous period, $m$, such that $Y_m = 0$); $A$ is the indicator for treatment; and $L$ is a vector of potential confounders at baseline.

We then used the predicted probabilities from these models, to estimate period-specific cumulative probabilities of being event-free for each individual, $i$, under each treatment strategy conditional on the individual's baseline confounders at, $L_i$. For individual $i$ and period $k$, we estimated the survival probability under treatment $a$ using the following formula:

$$\widehat{OM}_{i,k}^{a} = \prod_{m=1}^{k} \left\{ 1 - expit(\widehat{\beta}_0 + \widehat{\beta}_1 m + \widehat{\beta}_2 m^2 + \widehat{\beta}_3 a + \widehat{\beta}_4 am + \widehat{\beta}_5 am^2 + \widehat{\beta}_6^T L_i) \right\}.$$

We then standardized the survival probabilities at each time point to the empirical distribution of the baseline confounders:

$$\widehat{OM}_{k}^{a} = \frac{1}{n} \sum_{i=1}^{n} \widehat{OM}_{i,k}^{a},$$

where $n$ is the number of individuals in the study sample. The risk at time $k$, can then be estimated by taking one minus $\widehat{OM}_k^a$; risk differences and risk ratios can be calculated using the estimated risks (under each treatment). Finally, we used nonparametric bootstrapping with 200 samples to construct 95% confidence intervals.

#### Appendix 5 - Baseline characteristics of eligible individuals from an observational emulation of a target trial of thrombus aspiration vs. no thrombus aspiration, SWEDEHEART registry, 2007-2016, with unweighted and inverse probability weighted standardized mean differences

| | Thrombus aspiration | No thrombus aspiration | unweighted SMD | IP weighted SMD |
|---|---|---|---|---|
| **n** | 3462 | 14760 | | |
| **Age (yrs) (median [IQR])** | 66.0 [57.0, 74.0] | 68.0 [60.0, 77.0] | 0.190 | 0.043 |
| **Female (%)** | 887 (25.6) | 4422 (30.0) | 0.097 | 0.023 |
| **Hospital (%)** | | | 0.649 | 0.128 |
| Borås | 15 (0.4) | 143 (1.0) | | |
| Danderyd | 134 (3.9) | 363 (2.5) | | |
| Eskilstuna | 67 (1.9) | 398 (2.7) | | |
| Falun | 211 (6.1) | 638 (4.3) | | |
| Gävle | 333 (9.6) | 506 (3.4) | | |
| Halmstad | 13 (0.4) | 267 (1.8) | | |
| Helsingborg | 22 (0.6) | 90 (0.6) | | |
| Huddinge | 28 (0.8) | 131 (0.9) | | |
| Jönköping | 49 (1.4) | 679 (4.6) | | |
| Kalmar | 84 (2.4) | 566 (3.8) | | |
| Karlskrona | 165 (4.8) | 619 (4.2) | | |
| Karlstad | 65 (1.9) | 778 (5.3) | | |
| Karolinska Solna | 255 (7.4) | 1099 (7.4) | | |
| Kristianstad | 4 (0.1) | 144 (1.0) | | |
| Linköping | 251 (7.3) | 713 (4.8) | | |
| Lund | 785 (22.7) | 1929 (13.1) | | |
| Malmö | 32 (0.9) | 199 (1.3) | | |
| Sahlgrenska | 164 (4.7) | 1387 (9.4) | | |
| Skövde | 50 (1.4) | 487 (3.3) | | |
| St Görans | 42 (1.2) | 62 (0.4) | | |
| Sunderbyn | 19 (0.5) | 281 (1.9) | | |
| Sundsvall | 26 (0.8) | 214 (1.4) | | |
| SÖS | 170 (4.9) | 260 (1.8) | | |
| Trollhättan | 65 (1.9) | 328 (2.2) | | |
| Umeå | 33 (1.0) | 327 (2.2) | | |
| Uppsala | 160 (4.6) | 810 (5.5) | | |
| Västerås | 85 (2.5) | 283 (1.9) | | |
| Örebro | 125 (3.6) | 998 (6.8) | | |
| Östersund | 6 (0.2) | 47 (0.3) | | |
| Östra sjukhuset | 4 (0.1) | 14 (0.1) | | |
| **Stenosis class (%)** | | | 0.213 | 0.049 |
| A | 168 (4.9) | 928 (6.3) | | |
| B1 | 882 (25.5) | 4550 (30.8) | | |
| B2 | 1346 (38.9) | 6035 (40.9) | | |

| | Thrombus aspiration | No thrombus aspiration | unweighted SMD | IP weighted SMD |
|---|---|---|---|---|
| C | 1060 (30.6) | 3214 (21.8) | | |
| Other | 6 (0.2) | 33 (0.2) | | |
| **Stenosis in culprit artery (%)** | | | 0.441 | 0.151 |
| 50-69% | 34 (1.0) | 198 (1.3) | | |
| 70-89% | 118 (3.4) | 1081 (7.3) | | |
| 90-99% | 510 (14.7) | 4406 (29.9) | | |
| 100% | 2800 (80.9) | 9075 (61.5) | | |
| **Angiography finding (%)** | | | 0.164 | 0.032 |
| Normal | 2 (0.1) | 16 (0.1) | | |
| 1 vessel | 1957 (56.5) | 7260 (49.2) | | |
| 2 vessels | 931 (26.9) | 4271 (28.9) | | |
| 3 vessels | 450 (13.0) | 2537 (17.2) | | |
| Left main | 117 (3.4) | 659 (4.5) | | |
| Missing | 5 (0.1) | 17 (0.1) | | |
| **BMI (kg/m^2) (median [IQR])** | 26.0 [24.0, 29.0] | 26.0 [24.0, 29.0] | 0.014 | 0.005 |
| Missing (%) | 839 (24.2) | 3642 (24.7) | | |
| **Smoking status (%)** | | | 0.104 | 0.021 |
| Never | 1157 (33.4) | 5596 (37.9) | | |
| Ex smoker (> 1 month) | 957 (27.6) | 4045 (27.4) | | |
| Current smoker | 1038 (30.0) | 3999 (27.1) | | |
| Missing | 310 (9.0) | 1120 (7.6) | | |
| **Diabetes (%)** | 428 (12.4) | 2292 (15.5) | 0.091 | 0.024 |
| **Hyperlipidemia treatment (%)** | 724 (20.9) | 3327 (22.5) | 0.039 | 0.003 |
| **Hypertension treatment (%)** | 1340 (38.7) | 6628 (44.9) | 0.126 | 0.022 |
| **Previous myocardial infarction (%)** | 426 (12.3) | 1996 (13.5) | 0.036 | 0.016 |
| **Previous percutaneous coronary intervention (%)** | 368 (10.6) | 1563 (10.6) | 0.001 | 0.017 |
| **Previous coronary artery bypass grafting (%)** | 64 (1.8) | 337 (2.3) | 0.031 | 0.005 |
| **Thrombolysis (%)** | 16 (0.5) | 54 (0.4) | 0.015 | 0.007 |
| **Warfarin (%)** | 72 (2.1) | 303 (2.1) | 0.002 | 0.004 |
| **Aspirin (%)** | 3341 (96.5) | 14347 (97.2) | 0.040 | 0.009 |
| **Clopidogrel or ticlopidine (%)** | 2141 (61.8) | 6357 (43.1) | 0.383 | 0.031 |
| **Prasugrel (%)** | 118 (3.4) | 623 (4.2) | 0.042 | 0.028 |
| **Heparin (%)** | 2796 (80.8) | 12648 (85.7) | 0.132 | 0.018 |
| **Low-molecular weight heparin (%)** | 311 (9.0) | 883 (6.0) | 0.114 | 0.022 |
| **Bivalirudin (%)** | 1729 (49.9) | 7081 (48.0) | 0.039 | 0.021 |
| **Glycoprotein IIb/IIIa inhibitors (%)** | 1457 (42.1) | 3906 (26.5) | 0.334 | 0.076 |
| **Heart rate (median [IQR])** | 74.0 [61.0, 87.0] | 75.0 [63.0, 88.0] | 0.090 | 0.042 |
| Missing (%) | 245 (7.1) | 784 (5.3) | | |
| **Systolic blood pressure (median [IQR])** | 138.0 [120.0, 157.0] | 141.0 [125.0, 160.0] | 0.181 | 0.066 |
| Missing (%) | 257 (7.4) | 837 (5.7) | | |
| **Diastolic blood pressure (median [IQR])** | 80.0 [70.0, 95.0] | 84.0 [72.0, 96.0] | 0.088 | 0.033 |
| Missing (%) | 457 (13.2) | 1475 (10.0) | | |

#### Appendix 6 - Baseline characteristics of eligible individuals for an observational emulation of a target trial of thrombus aspiration vs. no thrombus aspiration, SWEDEHEART registry, stratified into 2007-2010 & 2013-2016

| | Before TASTE (Sep 07 to Jun 10) | | After TASTE (Mar 13 to Jan 16) | |
|---|---|---|---|---|
| | **Thrombus aspiration** | **No thrombus aspiration** | **Thrombus aspiration** | **No thrombus aspiration** |
| **n** | 2057 | 5602 | 1405 | 9158 |
| **Age (yrs) (median [IQR])** | 65.0 [58.0, 74.0] | 68.0 [60.0, 77.0] | 67.0 [57.0, 75.0] | 68.0 [60.0, 77.0] |
| **Female (%)** | 517 (25.1) | 1712 (30.6) | 370 (26.3) | 2710 (29.6) |
| **Hospital (%)** | | | | |
| Borås | 8 (0.4) | 55 (1.0) | 7 (0.5) | 88 (1.0) |
| Danderyd | 33 (1.6) | 46 (0.8) | 101 (7.2) | 317 (3.5) |
| Eskilstuna | 3 (0.1) | 4 (0.1) | 64 (4.6) | 394 (4.3) |
| Falun | 181 (8.8) | 256 (4.6) | 30 (2.1) | 382 (4.2) |
| Gävle | 193 (9.4) | 238 (4.2) | 140 (10.0) | 268 (2.9) |
| Halmstad | 5 (0.2) | 92 (1.6) | 8 (0.6) | 175 (1.9) |
| Helsingborg | 1 (0.0) | 0 (0.0) | 21 (1.5) | 90 (1.0) |
| Huddinge | 8 (0.4) | 49 (0.9) | 20 (1.4) | 82 (0.9) |
| Jönköping | 10 (0.5) | 214 (3.8) | 39 (2.8) | 465 (5.1) |
| Kalmar | 71 (3.5) | 299 (5.3) | 13 (0.9) | 267 (2.9) |
| Karlskrona | 115 (5.6) | 270 (4.8) | 50 (3.6) | 349 (3.8) |
| Karlstad | 43 (2.1) | 416 (7.4) | 22 (1.6) | 362 (4.0) |
| Karolinska Solna | 133 (6.5) | 503 (9.0) | 122 (8.7) | 596 (6.5) |
| Kristianstad | 0 (0.0) | 74 (1.3) | 4 (0.3) | 70 (0.8) |
| Linköping | 119 (5.8) | 256 (4.6) | 132 (9.4) | 457 (5.0) |
| Lund | 596 (29.0) | 708 (12.6) | 189 (13.5) | 1221 (13.3) |
| Malmö | 29 (1.4) | 173 (3.1) | 3 (0.2) | 26 (0.3) |
| Sahlgrenska | 74 (3.6) | 393 (7.0) | 90 (6.4) | 994 (10.9) |
| Skövde | 5 (0.2) | 188 (3.4) | 45 (3.2) | 299 (3.3) |
| St Görans | 18 (0.9) | 10 (0.2) | 24 (1.7) | 52 (0.6) |
| Sunderbyn | 3 (0.1) | 7 (0.1) | 16 (1.1) | 274 (3.0) |
| Sundsvall | 11 (0.5) | 44 (0.8) | 15 (1.1) | 170 (1.9) |
| SÖS | 71 (3.5) | 17 (0.3) | 99 (7.0) | 243 (2.7) |
| Trollhättan | 57 (2.8) | 166 (3.0) | 8 (0.6) | 162 (1.8) |
| Umeå | 20 (1.0) | 70 (1.2) | 13 (0.9) | 257 (2.8) |
| Uppsala | 100 (4.9) | 435 (7.8) | 60 (4.3) | 375 (4.1) |
| Västerås | 55 (2.7) | 25 (0.4) | 30 (2.1) | 258 (2.8) |
| Örebro | 95 (4.6) | 592 (10.6) | 30 (2.1) | 406 (4.4) |
| Östersund | 0 (0.0) | 0 (0.0) | 6 (0.4) | 47 (0.5) |
| Östra sjukhuset | 0 (0.0) | 2 (0.0) | 4 (0.3) | 12 (0.1) |
| **Stenosis class (%)** | | | | |
| A | 113 (5.5) | 373 (6.7) | 55 (3.9) | 555 (6.1) |
| B1 | 492 (23.9) | 1670 (29.8) | 390 (27.8) | 2880 (31.4) |

| | Before TASTE (Sep 07 to Jun 10) | | After TASTE (Mar 13 to Jan 16) | |
|---|---|---|---|---|
| | **Thrombus aspiration** | **No thrombus aspiration** | **Thrombus aspiration** | **No thrombus aspiration** |
| B2 | 775 (37.7) | 2220 (39.6) | 571 (40.6) | 3815 (41.7) |
| C | 675 (32.8) | 1314 (23.5) | 385 (27.4) | 1900 (20.7) |
| Other | 2 (0.1) | 25 (0.4) | 4 (0.3) | 8 (0.1) |
| **Stenosis in culprit artery (%)** | | | | |
| 50-69% | 19 (0.9) | 58 (1.0) | 15 (1.1) | 140 (1.5) |
| 70-89% | 62 (3.0) | 376 (6.7) | 56 (4.0) | 705 (7.7) |
| 90-99% | 318 (15.5) | 1661 (29.7) | 192 (13.7) | 2745 (30.0) |
| 100% | 1658 (80.6) | 3507 (62.6) | 1142 (81.3) | 5568 (60.8) |
| **Angiography finding (%)** | | | | |
| Normal | 2 (0.1) | 13 (0.2) | 0 (0.0) | 3 (0.0) |
| 1 vessel | 1134 (55.1) | 2593 (46.3) | 823 (58.6) | 4667 (51.0) |
| 2 vessels | 543 (26.4) | 1728 (30.8) | 388 (27.6) | 2543 (27.8) |
| 3 vessels | 307 (14.9) | 1007 (18.0) | 143 (10.2) | 1530 (16.7) |
| Left main | 70 (3.4) | 250 (4.5) | 47 (3.3) | 409 (4.5) |
| Missing | 1 (0.0) | 11 (0.2) | 4 (0.3) | 6 (0.1) |
| **BMI (kg/m^2) (median [IQR])** | 26.0 [24.0, 29.0] | 26.0 [24.0, 29.0] | 27.0 [24.0, 29.0] | 26.0 [24.0, 29.0] |
| Missing (%) | 480 (23.3) | 1567 (28.0) | 359 (25.6) | 2075 (22.7) |
| **Smoking status (%)** | | | | |
| Never | 664 (32.3) | 2121 (37.9) | 493 (35.1) | 3475 (37.9) |
| Ex smoker (> 1 month) | 563 (27.4) | 1440 (25.7) | 394 (28.0) | 2605 (28.4) |
| Current smoker | 641 (31.2) | 1579 (28.2) | 397 (28.3) | 2420 (26.4) |
| Missing | 189 (9.2) | 462 (8.2) | 121 (8.6) | 658 (7.2) |
| **Diabetes (%)** | 251 (12.2) | 802 (14.3) | 177 (12.6) | 1490 (16.3) |
| **Hyperlipidemia treatment (%)** | 404 (19.6) | 1195 (21.3) | 320 (22.8) | 2132 (23.3) |
| **Hypertension treatment (%)** | 777 (37.8) | 2296 (41.0) | 563 (40.1) | 4332 (47.3) |
| **Previous myocardial infarction (%)** | 256 (12.4) | 811 (14.5) | 170 (12.1) | 1185 (12.9) |
| **Previous percutaneous coronary intervention (%)** | 223 (10.8) | 609 (10.9) | 145 (10.3) | 954 (10.4) |
| **Previous coronary artery bypass grafting (%)** | 31 (1.5) | 133 (2.4) | 33 (2.3) | 204 (2.2) |
| **Thrombolysis (%)** | 10 (0.5) | 19 (0.3) | 6 (0.4) | 35 (0.4) |
| **Warfarin (%)** | 30 (1.5) | 106 (1.9) | 42 (3.0) | 197 (2.2) |
| **Aspirin (%)** | 1980 (96.3) | 5391 (96.2) | 1361 (96.9) | 8956 (97.8) |
| **Clopidogrel or ticlopidine (%)** | 1903 (92.5) | 5247 (93.7) | 238 (16.9) | 1110 (12.1) |
| **Prasugrel (%)** | 75 (3.6) | 49 (0.9) | 43 (3.1) | 574 (6.3) |
| **Heparin (%)** | 1506 (73.2) | 4027 (71.9) | 1290 (91.8) | 8621 (94.1) |
| **Low-molecular weight heparin (%)** | 281 (13.7) | 781 (13.9) | 30 (2.1) | 102 (1.1) |
| **Bivalirudin (%)** | 836 (40.6) | 1790 (32.0) | 893 (63.6) | 5291 (57.8) |
| **Glycoprotein IIb/IIIa inhibitors (%)** | 1119 (54.4) | 2942 (52.5) | 338 (24.1) | 964 (10.5) |
| **Heart rate (median [IQR])** | 72.0 [60.0, 85.0] | 74.0 [62.0, 87.0] | 75.0 [62.0, 88.0] | 75.0 [64.0, 89.0] |
| Missing (%) | 216 (10.5) | 595 (10.6) | 29 (2.1) | 189 (2.1) |
| **Systolic blood pressure (median [IQR])** | 138.0 [120.0, 156.0] | 140.0 [122.0, 160.0] | 139.5 [120.0, 158.0] | 144.0 [125.0, 162.2] |
| Missing (%) | 226 (11.0) | 647 (11.5) | 31 (2.2) | 190 (2.1) |

| | Before TASTE (Sep 07 to Jun 10) | | After TASTE (Mar 13 to Jan 16) | |
|---|---|---|---|---|
| | **Thrombus aspiration** | **No thrombus aspiration** | **Thrombus aspiration** | **No thrombus aspiration** |
| **Diastolic blood pressure (median [IQR])** | 80.0 [70.0, 95.0] | 80.0 [70.0, 95.0] | 82.5 [70.0, 95.0] | 85.0 [74.0, 97.0] |
| Missing (%) | 316 (15.4) | 803 (14.3) | 141 (10.0) | 672 (7.3) |

#### Appendix 7 - Estimated 1-year risk, risk difference, and risk ratios from an observational emulation of a target trial of thrombus aspiration vs. no thrombus aspiration, SWEDEHEART registry, stratified into 2007-2010 & 2013-2016

| | | Risk (%, 95% CI) | | | |
|---|---|---|---|---|---|
| **Outcome** | **Model** | **Thrombus aspiration** | **No thrombus aspiration** | **Risk difference (%, 95% CI)** | **Risk ratio (95% CI)** |
| **Before TASTE** | | | | | |
| Death | Age and sex adjusted | 7.7 (6.1, 9.3) | 7.3 (6.3, 8.3) | 0.4 (-1.4, 2.2) | 1.05 (0.89, 1.25) |
| | Full adjustment* | 7.3 (5.8, 8.8) | 7.5 (6.5, 8.4) | -0.2 (-2.0, 1.6) | 0.97 (0.81, 1.17) |
| Myocardial infarction | Age and sex adjusted | 4.2 (3.0, 5.4) | 5.1 (4.3, 5.9) | -0.9 (-2.3, 0.6) | 0.83 (0.66, 1.03) |
| | Full adjustment* | 4.3 (3.1, 5.6) | 5.0 (4.3, 5.8) | -0.7 (-2.3, 0.8) | 0.85 (0.68, 1.08) |
| **After TASTE** | | | | | |
| Death | Age and sex adjusted | 9.4 (7.4, 11.4) | 7.2 (6.4, 8.0) | 2.2 ( 0.0, 4.4) | 1.31 (1.10, 1.56) |
| | Full adjustment* | 8.5 (6.7, 10.4) | 7.3 (6.6, 8.1) | 1.2 (-0.7, 3.2) | 1.17 (0.98, 1.39) |
| Myocardial infarction | Age and sex adjusted | 3.8 (2.4, 5.2) | 3.4 (2.9, 3.9) | 0.4 (-1.0, 1.9) | 1.12 (0.86, 1.47) |
| | Full adjustment* | 3.6 (2.2, 5.1) | 3.4 (2.9, 3.9) | 0.2 (-1.3, 1.8) | 1.06 (0.79, 1.44) |

*Adjusted at baseline for: age, sex, hospital, diabetes, body mass index, smoking, hyperlipidemia, hypertension, previous infarction, previous percutaneous coronary intervention, previous coronary artery bypass graft, stenosis class, proportion stenosis, angiography finding, heart rate, systolic blood pressure, diastolic blood pressure, thrombolysis, warfarin, aspirin, clopidogrel, prasugrel, heparin, low molecular weight heparin, bivalirudin, and Gp2b3a inhibitors

#### Appendix 8 - Average 1-year hazard ratio from TASTE and estimated 1-year risk ratios from an observational emulation of a target trial of thrombus aspiration vs. no thrombus aspiration, SWEDEHEART registry, stratified into 2007-2010 & 2013-2016

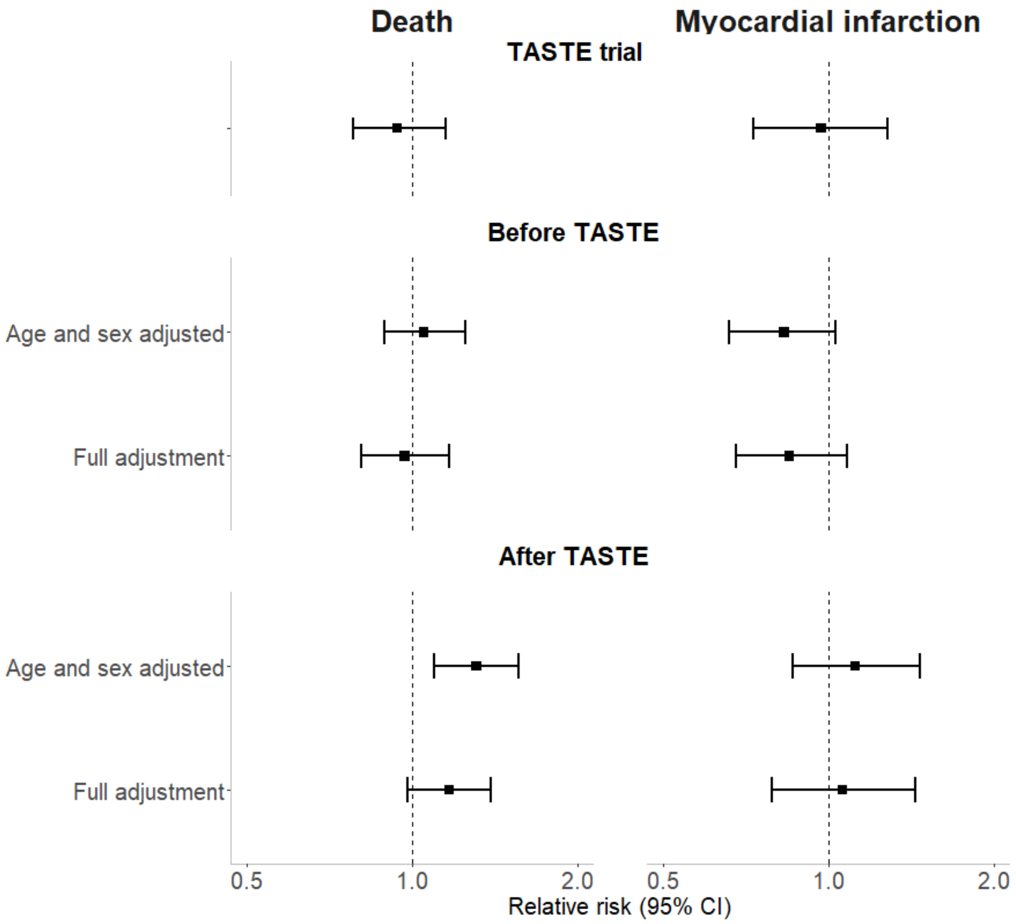

**Appendix 9 - Sensitivity analysis - Estimated 1-year risk, risk difference, and risk ratios from an observational emulation of a target trial of thrombus aspiration vs. no thrombus aspiration, SWEDEHEART register, 2007-2016, without application of the 50% minimum stenosis eligibility criterion**

| | Risk (%, 95% CI)* | | | |
|---|---|---|---|---|
| **Outcome** | **Thrombus aspiration** | **No thrombus aspiration** | **Risk difference (%, 95% CI)*** | **Risk ratio (95% CI)*** |
| Death | 8.1 (7.0, 9.3) | 7.3 (6.8, 7.9) | 0.8 (-0.5, 2.0) | 1.11 (0.98, 1.24) |
| Myocardial infarction | 4.0 (3.1, 4.9) | 4.3 (3.9, 4.8) | -0.3 (-1.3, 0.7) | 0.94 (0.79, 1.11) |

*Adjusted at baseline for: age, sex, hospital, diabetes, body mass index, smoking, hyperlipidemia, hypertension, previous infarction, previous percutaneous coronary intervention, previous coronary artery bypass graft, stenosis class, proportion stenosis, angiography finding, heart rate, systolic blood pressure, diastolic blood pressure, thrombolysis, warfarin, aspirin, clopidogrel, prasugrel, heparin, low molecular weight heparin, bivalirudin, and Gp2b3a inhibitors

#### Appendix 10 - Sensitivity analysis - Estimated 1-year risk, risk difference, and risk ratios from an observational emulation of a target trial of thrombus aspiration vs. no thrombus aspiration, SWEDEHEART register, 2007-2016, adjusted for age and sex

| | Risk (%, 95% CI) | | | |
|---|---|---|---|---|
| **Outcome** | **Thrombus aspiration*** | **No thrombus aspiration*** | **Risk difference (%, 95% CI)*** | **Risk ratio (95% CI)*** |
| Death | 8.5 (7.2, 9.8) | 7.2 (6.6, 7.8) | 1.3 (-0.1, 2.7) | 1.18 (1.04, 1.34) |
| Myocardial infarction | 4.1 (3.0, 5.1) | 4.0 (3.6, 4.5) | 0.0 (-1.1, 1.2) | 1.01 (0.83, 1.22) |

*Adjusted at baseline for: age and sex

**Appendix 11 - Sensitivity analysis - Difference in risk differences between age and sex adjusted, and fully adjusted models from an observational emulation of a target trial of thrombus aspiration vs. no thrombus aspiration, SWEDEHEART register, 2007-2016**

| Outcome | Difference in risk difference |
|---|---|
| Death | -0.6 (-1.4, 0.1) |
| Myocardial infarction | -0.2 (-0.6, 0.2) |

#### Appendix 12 - Sensitivity analysis - Estimated 1-year risk, risk difference, and risk ratios from an observational emulation of a target trial of thrombus aspiration vs. no thrombus aspiration, SWEDEHEART register, 2007-2016, with individuals censored at death

| | Risk (%, 95% CI)* | | | |
|---|---|---|---|---|
| **Outcome** | **Thrombus aspiration** | **No thrombus aspiration** | **Risk difference (%, 95% CI)*** | **Risk ratio (95% CI)*** |
| Myocardial infarction | 4.1 (3.2,5.1) | 4.3 (3.8,4.8) | -0.2 (-1.2,0.8) | 0.95 (0.80,1.14) |

*Adjusted at baseline for: age, sex, hospital, diabetes, body mass index, smoking, hyperlipidemia, hypertension, previous infarction, previous percutaneous coronary intervention, previous coronary artery bypass graft, stenosis class, proportion stenosis, angiography finding, heart rate, systolic blood pressure, diastolic blood pressure, thrombolysis, warfarin, aspirin, clopidogrel, prasugrel, heparin, low molecular weight heparin, bivalirudin, and Gp2b3a inhibitors

#### Appendix 13 - Sensitivity analysis - Estimated 1-year risk, risk difference, and risk ratios from an observational emulation of a target trial of thrombus aspiration vs. no thrombus aspiration, SWEDEHEART register, 2007-2016, with a complete case analysis

| | Risk (%, 95% CI)* | | | |
|---|---|---|---|---|
| **Outcome** | **Thrombus aspiration** | **No thrombus aspiration** | **Risk difference (%, 95% CI)*** | **Risk ratio (95% CI)*** |
| Death | 6.5 (4.7,8.3) | 5.6 (4.7,6.4) | 0.9 (-1.1,2.9) | 1.16 (0.92,1.46) |
| Myocardial infarction | 4.2 (2.7,5.7) | 4.4 (3.5,5.2) | -0.2 (-2.0,1.6) | 0.96 (0.71,1.28) |

*Adjusted at baseline for: age, sex, hospital, diabetes, body mass index, smoking, hyperlipidemia, hypertension, previous infarction, previous percutaneous coronary intervention, previous coronary artery bypass graft, stenosis class, proportion stenosis, angiography finding, heart rate, systolic blood pressure, diastolic blood pressure, thrombolysis, warfarin, aspirin, clopidogrel, prasugrel, heparin, low molecular weight heparin, bivalirudin, and Gp2b3a inhibitors

#### Appendix 14 - Sensitivity analysis - Estimated 1-year risk, risk difference, and risk ratios from an observational emulation of a target trial of thrombus aspiration vs. no thrombus aspiration, SWEDEHEART register, 2007-2016, with alternative myocardial infarction definition

| | Risk (%, 95% CI)* | | | |
|---|---|---|---|---|
| **Outcome** | **Thrombus aspiration** | **No thrombus aspiration** | **Risk difference (%, 95% CI)*** | **Risk ratio (95% CI)*** |
| Myocardial infarction | 3.8 (2.8, 4.8) | 3.8 (3.4, 4.3) | 0.0 (-1.2, 1.1) | 0.99 (0.81, 1.21) |

*Adjusted at baseline for: age, sex, hospital, diabetes, body mass index, smoking, hyperlipidemia, hypertension, previous infarction, previous percutaneous coronary intervention, previous coronary artery bypass graft, stenosis class, proportion stenosis, angiography finding, heart rate, systolic blood pressure, diastolic blood pressure, thrombolysis, warfarin, aspirin, clopidogrel, prasugrel, heparin, low molecular weight heparin, bivalirudin, and Gp2b3a inhibitors

#### Appendix 15 - Sensitivity analysis - Estimated 1-year risk, risk difference, and risk ratios from an observational emulation of a target trial of thrombus aspiration vs. no thrombus aspiration, SWEDEHEART register, 2013-2016, additionally adjusted for Killip class

| | **Risk (%, 95% CI)*** | | | |
|---|---|---|---|---|
| **Outcome** | **Thrombus aspiration** | **No thrombus aspiration** | **Risk difference (%, 95% CI)*** | **Risk ratio (95% CI)*** |
| Death | 8.4 (6.6, 10.1) | 7.3 (6.6, 8.1) | 1.0 (-0.9, 2.9) | 1.14 (0.96, 1.35) |
| Myocardial infarction | 3.6 (2.3, 5.0) | 3.4 (2.9, 3.9) | 0.2 (-1.2, 1.7) | 1.06 (0.80, 1.40) |

*Adjusted at baseline for: killip class, age, sex, hospital, diabetes, body mass index, smoking, hyperlipidemia, hypertension, previous infarction, previous percutaneous coronary intervention, previous coronary artery bypass graft, stenosis class, proportion stenosis, angiography finding, heart rate, systolic blood pressure, diastolic blood pressure, thrombolysis, warfarin, aspirin, clopidogrel, prasugrel, heparin, low molecular weight heparin, bivalirudin, and Gp2b3a inhibitors

#### Appendix 16 - Sensitivity analysis - Estimated 1-year risk, risk difference, and risk ratios from an observational emulation of a target trial of thrombus aspiration vs. no thrombus aspiration, SWEDEHEART register, 2013-2016, additionally adjusted for time period (before or after TASTE)

| | Risk (%, 95% CI)* | | | |
|---|---|---|---|---|
| **Outcome** | **Thrombus aspiration** | **No thrombus aspiration** | **Risk difference (%, 95% CI)*** | **Risk ratio (95% CI)*** |
| Death | 8.0 (6.7,9.3) | 7.3 (6.8,7.9) | 0.7 (-0.7,2.1) | 1.09 (0.96,1.24) |
| Myocardial infarction | 3.9 (2.9,4.9) | 4.1 (3.6,4.5) | -0.2 (-1.3,0.9) | 0.95 (0.78,1.16) |

*Adjusted at baseline for: period, age, sex, hospital, diabetes, body mass index, smoking, hyperlipidemia, hypertension, previous infarction, previous percutaneous coronary intervention, previous coronary artery bypass graft, stenosis class, proportion stenosis, angiography finding, heart rate, systolic blood pressure, diastolic blood pressure, thrombolysis, warfarin, aspirin, clopidogrel, prasugrel, heparin, low molecular weight heparin, bivalirudin, and Gp2b3a inhibitors

#### Appendix 17 - Sensitivity analysis - Estimated 1-year risks from an observational emulation of a target trial for thrombus aspiration and no thrombus aspiration, SWEDEHEART register, 2013-2016, when data are stratified by exposure ad outcome models are fit in each arm

| | Risk (%, 95% CI)* | |
|---|---|---|
| **Outcome** | **Thrombus aspiration** | **No thrombus aspiration** |
| Death | 7.5 (6.9,8.0) | 7.5 (6.3,8.6) |
| Myocardial infarction | 4.1 (3.6,4.5) | 4.0 (3.0,4.9) |

*Adjusted at baseline for: age, sex, hospital, diabetes, body mass index, smoking, hyperlipidemia, hypertension, previous infarction, previous percutaneous coronary intervention, previous coronary artery bypass graft, stenosis class, proportion stenosis, angiography finding, heart rate, systolic blood pressure, diastolic blood pressure, thrombolysis, warfarin, aspirin, clopidogrel, prasugrel, heparin, low molecular weight heparin, bivalirudin, and Gp2b3a inhibitors

**Appendix 18 - Sensitivity analysis - Estimated 1-year risk, risk difference, and risk ratios from an observational emulation of a target trial of thrombus aspiration vs. no thrombus aspiration, SWEDEHEART register, 2013-2016, adjusted for baseline confounding using inverse probability weighting**

| | Risk (%, 95% CI)* | | | |
|---|---|---|---|---|
| **Outcome** | **Thrombus aspiration** | **No thrombus aspiration** | **Risk difference (%, 95% CI)*** | **Risk ratio (95% CI)*** |
| Death | 8.3 (7.3,9.2) | 7.4 (7.1,7.7) | 0.9 (-0.1,1.9) | 1.12 (0.99,1.26) |
| Myocardial infarction | 3.7 (3.1,4.3) | 4.1 (3.9,4.3) | -0.4 (-1.1,0.2) | 0.90 (0.75,1.07) |

*Adjusted at baseline for: period, age, sex, hospital, diabetes, body mass index, smoking, hyperlipidemia, hypertension, previous infarction, previous percutaneous coronary intervention, previous coronary artery bypass graft, stenosis class, proportion stenosis, angiography finding, heart rate, systolic blood pressure, diastolic blood pressure, thrombolysis, warfarin, aspirin, clopidogrel, prasugrel, heparin, low molecular weight heparin, bivalirudin, and Gp2b3a inhibitors
